# Predictors of Visual Acuity, Intraocular Pressure, and Pain in Neovascular Glaucoma: A Mixed-Effects Model and Machine Learning Cohort Analysis

**DOI:** 10.64898/2026.08.11.26360163

**Authors:** Gilbert Simons, Mikael von Fersen, Alexandra Dahlberg, Ville Vartiainen, Paula Summanen, Mika Harju

**Affiliations:** Faculty of Medicine, University of Helsinki, Helsinki, Finland; Department of Ophthalmology, Helsinki University Hospital, Helsinki, Finland; Heart and Lung Centre, Helsinki University Hospital, Helsinki, Finland

**Keywords:** Glaucoma, Neovascular, Visual Acuity, Intraocular Pressure, Machine Learning, Treatment Outcome

## Abstract

**Background/Aims:** Neovascular glaucoma (NVG) is a severe, secondary glaucoma. This study aimed to identify factors associated with vision, intraocular pressure (IOP), and ocular pain outcomes.

**Methods:** The cohort included all patients diagnosed with NVG during 2008–2024 at Helsinki University Hospital, Finland. Linear mixed-effects models used pre-specified covariates, whereas machine learning was given the full longitudinal data with biomicroscopic findings as an exploratory approach.

**Results:** 626 patients were analysed. Worse baseline vision and a closed angle were associated with worse follow-up vision. Treatments were associated with lower IOP and less pain rather than better vision. Age, sex and comorbidity were largely not associated with the outcomes. Glaucoma drainage devices showed the greatest initial IOP reduction (−10.2 mmHg, 95% confidence interval, CI −11.9 to −8.6 mmHg), followed by transscleral cyclophotocoagulation (TSCPC, −4.7 mmHg, 95% CI −5.8 to −3.7 mmHg) and peripheral retinal cryotherapy (−2.2 mmHg, 95% CI −3.1 to −1.4 mmHg). TSCPC and cryotherapy were also associated with reduced pain (odds ratio 0.51 and 0.46). Pan-retinal photocoagulation and anti-VEGF showed smaller IOP reductions, with a pain reduction for pan-retinal photocoagulation only. Both methods agreed, and machine learning added no novel clinical findings.

**Conclusions:** Vision in this cohort was largely set by the state of the eye at diagnosis. IOP control and pain relief therefore remain realistic goals even when sight cannot be saved. Peripheral retinal cryotherapy stood out, linked to both lower IOP and less pain, seldom reported in NVG. These associations from a large, unselected cohort identify treatments worth comparing prospectively.

**Key Messages:** *What is already known on this topic:* Neovascular glaucoma has a poor visual prognosis. Treatments have mostly been assessed in operated patients as surgical success or failure, rather than by their associations with vision, IOP and pain across the disease course.

*What this study adds:* Treatments were associated mainly with lower intraocular pressure and less pain, not with better vision, which baseline severity largely determined. Peripheral retinal cryotherapy was associated with lower IOP and less pain, an effect seldom reported.

*How this study might affect research, practice or policy:* Intraocular pressure control and pain relief remain achievable goals, whereas visual prognosis is largely set at presentation.

**Précis:** In 626 neovascular glaucoma patients followed over 12,000 visits, baseline severity shaped visual prognosis. Treatments were associated with lower intraocular pressure and less ocular pain but rarely better vision. Mixed-effects models and machine learning agreed.

## Introduction

Neovascular glaucoma (NVG) is a severe, secondary glaucoma. It develops in response to ischaemia, commonly caused by central retinal vein occlusion (CRVO), diabetic retinopathy (DR), central retinal artery occlusion (CRAO), and ocular ischaemic syndrome (OIS)^1^ ^2^. Treating the disease involves both managing the underlying ischaemia and lowering intraocular pressure (IOP)^3^.

The ischaemia is often treated with pan-retinal photocoagulation (PRP), peripheral retinal cryotherapy, and anti-VEGF injections (VEGF: vascular endothelial growth factor)^3^. Separately, the glaucoma is addressed with topical and systemic glaucoma medications, as well as surgical approaches, such as transscleral cyclophotocoagulation (TSCPC) and glaucoma drainage devices (GDD)^3^.

Best-corrected visual acuity (BCVA) outcomes are often poor even when IOP is lowered^4^ ^5^. Patients commonly present with ocular pain that frequently resolves during follow-up^6^. Prior work has identified factors associated with these outcomes, mostly baseline factors linked to discrete events such as no light perception (NLP) or surgical failure^7–9^. However, earlier studies were limited in size, which prevented separate analysis by aetiology, and focused primarily on surgical interventions^5^ ^7–10^. Because most of these studies drew only on baseline data or on first and last visits, how treatments relate to the clinical course of BCVA, IOP, and pain has rarely been examined. A study applying machine learning (ML) to NVG have likewise relied only on baseline markers^11^. This study aims to analyse the full clinical course of all NVG patients treated at HUS Helsinki University Hospital between 2008 and 2024. We use linear mixed-effects models (LMMs) and ML to identify factors associated with BCVA, IOP, and pain outcomes.

## Methods

### Cohort

The cohort included all patients at HUS Helsinki University Hospital, Finland, diagnosed with NVG during 2008–2024. HUS is the only tertiary ophthalmological centre in its catchment area covering 32% of the Finnish population in 2024^12^. Patients were identified from electronic medical records (EMRs). Those with ICD-10 code H40.5 and a mention of NVG in at least one EMR were included. Additional inclusion criteria were a visit with IOP ≥25 mmHg with neovascularisation of the iris and/or anterior chamber angle. The date when inclusion criteria were first met was considered the time of diagnosis. Only patients followed primarily at HUS were included, with at least two visits from different dates. The study used the full eligible cohort with no formal sample-size calculation. The cohort was the same as described previously^13^. Reporting followed STROBE^14^.

### Included data

Baseline data included prior treatments, prior ocular, and prior systemic diseases. Missing baseline values were taken from the nearest visit within 30 days before or after diagnosis. Collected visit data comprised BCVA, IOP, pain, biomicroscopic findings, optical coherence tomography measurements, and treatments. BCVA was converted to the logarithm of the minimum angle of resolution (logMAR)^15^. Counting fingers, hand motion, light perception, and NLP were assigned values of 2.0, 2.3, 2.7, and 3.0^16^ ^17^, with counting fingers and hand motion adjusted by ±0.1 logMAR according to recorded distance. IOP was measured by Goldmann applanation tonometry or iCare rebound tonometry (Icare Finland Oy, Vantaa, Finland)^18^. Pain was categorised as none, present or severe. At each visit we extracted biomicroscopic findings covering the signs of neovascular activity and severity: corneal status, iris and angle neovascularisation, anterior chamber inflammation, peripheral anterior synechiae (PAS), and the state of the lens, vitreous, and retina. A closed angle was defined as Shaffer grade 0 or PAS over 270 degrees. Full variable grading scales and definitions, including Charlson Comorbidity Index (CCI)^19^, are available in Supplementary Methods.

Visits occurred at clinical discretion with no fixed follow-up schedule. A visit was included when it recorded any new information. Repeated BCVA, IOP, and pain measurements qualified even when numerically identical to the previous visit. Patients were not excluded for intermittent missing measurements. Follow-up ended at the last recorded ophthalmological visit. Some patients were referred to outsourced services for follow-up, and their records became available only from July 2022 onward.

### Treatments

PRP was the primary treatment for retinal ischaemia. Peripheral retinal cryotherapy was applied with or without direct visualisation, avoiding the 3 and 9 o’clock areas, often during the same periocular anaesthesia as TSCPC. Anti-VEGF agents used were aflibercept, ranibizumab, faricimab, and off-label bevacizumab. Their minimum dosing intervals were 56, 28, 56, and 28 days, respectively^3^ ^20–23^.

Medications included prostaglandins, beta blockers, topical and systemic carbonic anhydrase inhibitors, alpha agonists, parasympathomimetics, Rho-kinase inhibitors, topical cortisone, and anticholinergics. Medications were treated as individual binary values, carried forward with biomicroscopy findings to following visits until an update was recorded.

TSCPC was almost exclusively applied 180 degrees with 18–20 applications at a time. A slow coagulation technique^24^ was utilised using an effect of 430–600 mW with 8–10 s applications. The first application targeted inferiorly, the second nasally, and the third inferotemporally and superonasally, always leaving one quadrant untreated. GDDs used were Ahmed valves (New World Medical, Rancho Cucamonga, CA, USA), Baerveldt implants (Johnson & Johnson Surgical Vision, Santa Ana, CA, USA), and Paul implants (Advanced Ophthalmic Innovations, Singapore). Filtering surgery (trabeculectomy and non-penetrating deep sclerectomy) after diagnosis was not included due to rarity (n=6). Endarterectomy and vitrectomy were included.

### Statistical analysis

The primary outcomes were BCVA and IOP, with pain as secondary. Claude Opus 4 (Anthropic, San Francisco, CA, USA)^25^ assisted in drafting and debugging the analysis code, with all output verified against the source data.

### Linear mixed-effects models

LMMs were fitted to estimate associations of covariates with BCVA and IOP using Python (version 3.12.10, Python Software Foundation, Wilmington, DE, USA)^26^. A generalised linear mixed-effects model (GLMM) was fitted for pain using R Statistical Software (version 4.5.0, R Core Team 2026, Vienna, Austria)^27^. A random intercept and a random slope on time since diagnosis were fitted for each patient in the BCVA and IOP models. The pain GLMM also attempted a random slope but did not converge, so a random intercept only was retained. Time was modelled using a restricted cubic spline on days since diagnosis (Harrell three-knot, knots at 30, 180 and 730 days) to match potential non-linear trajectories^28^.

Continuous covariates were centred at clinical reference values and scaled (age per decade, visual acuity per logMAR unit, IOP per 10 mmHg). Categorical covariates used defined reference levels (CRVO, female sex, intraocular lens). Missing baseline values were retained through paired missing-data indicators. The covariate set for the LMMs was prespecified based on clinical relevance and data completeness, and no data-driven variable selection was used for these models. Treatments entered the models by mechanism. Cumulative-dose terms counted exposure. Step terms marked the immediate effect after a procedure. Time-since slopes captured change per year following a procedure. Anti-VEGF entered as a binary indicator, on for a fixed period after each injection and reset by each new injection, with the period set to each agent’s minimum dosing interval. Endarterectomy entered as a main effect and an interaction with OIS. All available visits were used. The BCVA model excluded eyes at NLP at baseline and censored at the first NLP visit. IOP used a combined measure of Goldmann applanation tonometry with rebound as fallback. Pain was binarised as any pain versus none. Robust sensitivity analyses were constructed for BCVA, IOP and pain (Supplementary Methods).

Restricted maximum likelihood was the estimation method for the LMMs and maximum likelihood (Laplace approximation) for the GLMM. BCVA and IOP were reported as coefficients and pain as odds ratios (OR) with 95% confidence intervals (CI; Wald). Two-sided *p*-values were reported, with *p*<0.05 considered nominally statistically significant.

### Machine learning

ML served as an exploratory complement to the LMM, identifying features associated with BCVA and IOP across the full recorded feature set. Post-diagnosis biomicroscopy findings were included, which the LMM omitted as potential mediators. Pain was not modelled by ML, as the binary, secondary outcome would have required a separate classification pipeline. Gradient-boosted trees were used because they capture non-linearities and interactions without pre-specification. eXtreme Gradient Boosting (XGBoost, version 2.0.3, Python)^29^ was selected for its prior use in glaucoma prediction studies^30–32^. Unlike the LMMs, this analysis treated each visit as independent, since XGBoost cannot model within-eye correlation. The model included the previous visit’s BCVA and IOP features, and analyses were run with and without these features to distinguish incremental from overall associations. All visits were used up to the last recorded visit or structural eye loss (enucleation, evisceration, phthisis, shell prosthesis, or alcohol injection). XGBoost handled missing values natively. Hyperparameters were pre-specified at conservative values rather than tuned, to prevent unstable combinations sometimes produced by formal tuning on modestly sized clinical datasets^30–33^. A learning rate of 0.05, max depth of 4 and 500 estimators were used (Supplementary Table 1). A sensitivity analysis varied max depth and learning rate.

Model performance was estimated using 5-fold cross-validation with patient-level grouping. All patients were retained in cross-validation to preserve power for rare treatment subgroups. Overfitting was assessed from the gap between training and validation root mean square error (RMSE), computed per visit and summarised across the five folds. To assess the contribution of each feature group, models were refitted excluding biomicroscopic features and using baseline features alone. Shapley Additive exPlanations (SHAP, version 0.46.0, Python) were implemented to characterise features driving predictions^34^. SHAP values for visualisation were computed from a model refitted on all patients, with cross-validation providing the generalisation estimate. Features whose ranking shifted by more than two positions across folds were considered unstable. We also examined interactions between features, mainly whether a treatment’s association with BCVA or IOP differed by the type of eye and repeated the treatment analysis with every patient weighted equally. Reporting followed TRIPOD+AI with applicability annotated per item^35^ (Supplementary Table 2).

### Study approval

The study was conducted under an institutional research permit from HUS (HUS/20/2025) and followed the Declaration of Helsinki. As a registry-based study, it did not require review by a medical research ethics committee or informed consent under Finnish legislation.

## Results

The cohort comprised 626 patients, with a median follow-up of 24 months (Table 1).

**Table 1.** Patient demographics, aetiologies, systemic and ocular history.

| Cohort characteristics | Baseline |
| --- | --- |
| Total patients, n | 626 |
| Age at diagnosis, years (IQR) | 76 (66–84) |
| Follow-up, months (IQR) | 24 (9–58) |
| Visits per patient (IQR) | 14 (9–22) |
| **Sex** |  |
| Male | 352 (56%) |
| Female | 274 (44%) |
| **Affected eye** |  |
| OD | 317 (51%) |
| OS | 309 (49%) |
| Bilateral neovascular glaucoma | 28 (4%) |
| **Aetiology** |  |
| Central retinal vein occlusion | 283 (45%) |
| Diabetic retinopathy | 90 (14%) |
| Central retinal artery occlusion | 66 (11%) |
| Ocular ischaemic syndrome | 49 (8%) |
| Other | 138 (22%) |
| **Systemic comorbidities** |  |
| Diabetes mellitus | 266 (42%) |
| Hypertension | 382 (61%) |
| Dyslipidaemia | 198 (32%) |
| Peripheral artery disease | 82 (13%) |
| **Prior glaucoma** |  |
| Glaucoma | 174 (28%) |
| Primary open-angle glaucoma | 79 (13%) |
| Exfoliation glaucoma | 78 (12%) |
| Normal-tension glaucoma | 4 (1%) |
| Other glaucoma subtype | 13 (2%) |
| **Prior ocular treatment<sup>1</sup>** |  |
| Anti-VEGF injections | 71 (11%) |
| Scatter retinal photocoagulation | 94 (15%) |
| Vitrectomy | 62 (10%) |
| Laser iridotomy | 27 (4%) |
| Selective laser trabeculoplasty | 17 (3%) |
| Glaucoma filtration surgery | 15 (2%) |
| Peripheral retinal cryotherapy | 5 (1%) |

<sup>1</sup>Any procedure or injection recorded before the date of NVG diagnosis. Prior ocular treatments are reported as presence or absence only; quantitative details, such as scatter retinal photocoagulation spot counts, were not captured. Scatter retinal photocoagulation includes panretinal photocoagulation and less extensive retinal laser treatments. IQR, interquartile range; VEGF, vascular endothelial growth factor.

### Linear mixed-effects model

The BCVA model included 509 patients (excluding eyes at NLP at baseline and missing baseline BCVA), the IOP model 626, and the pain model 621. Among the baseline factors, DR was associated with better BCVA than CRVO. Worse baseline BCVA was associated with worse vision and higher IOP across follow-up. A closed angle was associated with worse vision only, and higher baseline IOP with higher subsequent IOP. Higher age was associated with less pain. Sex and CCI were not statistically significant in any model.

PRP, per 1000 cumulative spots, was not associated with BCVA, but was related to lower IOP and lower odds of pain. Peripheral retinal cryotherapy was associated with better vision, lower IOP, and lower odds of pain. Anti-VEGF injections showed lower IOP in their active window but were not linked to BCVA or pain.

A higher number of glaucoma medications was associated with worse BCVA, higher IOP, and greater odds of pain. The first TSCPC session was associated with a step reduction in IOP and lower odds of pain, and each further 19 applications (cohort median) with a smaller additional reduction. The per-year term remained negative, indicating continued reduction after treatment. Pain odds ratio, by contrast, increased over time, exceeding 1. GDDs were associated with initially worse vision but lower IOP. The effect per year after GDD insertion reversed to better vision but was not statistically significant for IOP. GDDs did not reach statistical significance for pain. In OIS eyes, endarterectomy was associated with higher IOP and greater odds of pain. The difference from other aetiologies was significant for BCVA only. See Table 2 and Figure 1 for full LMM results.

**Figure 1.**
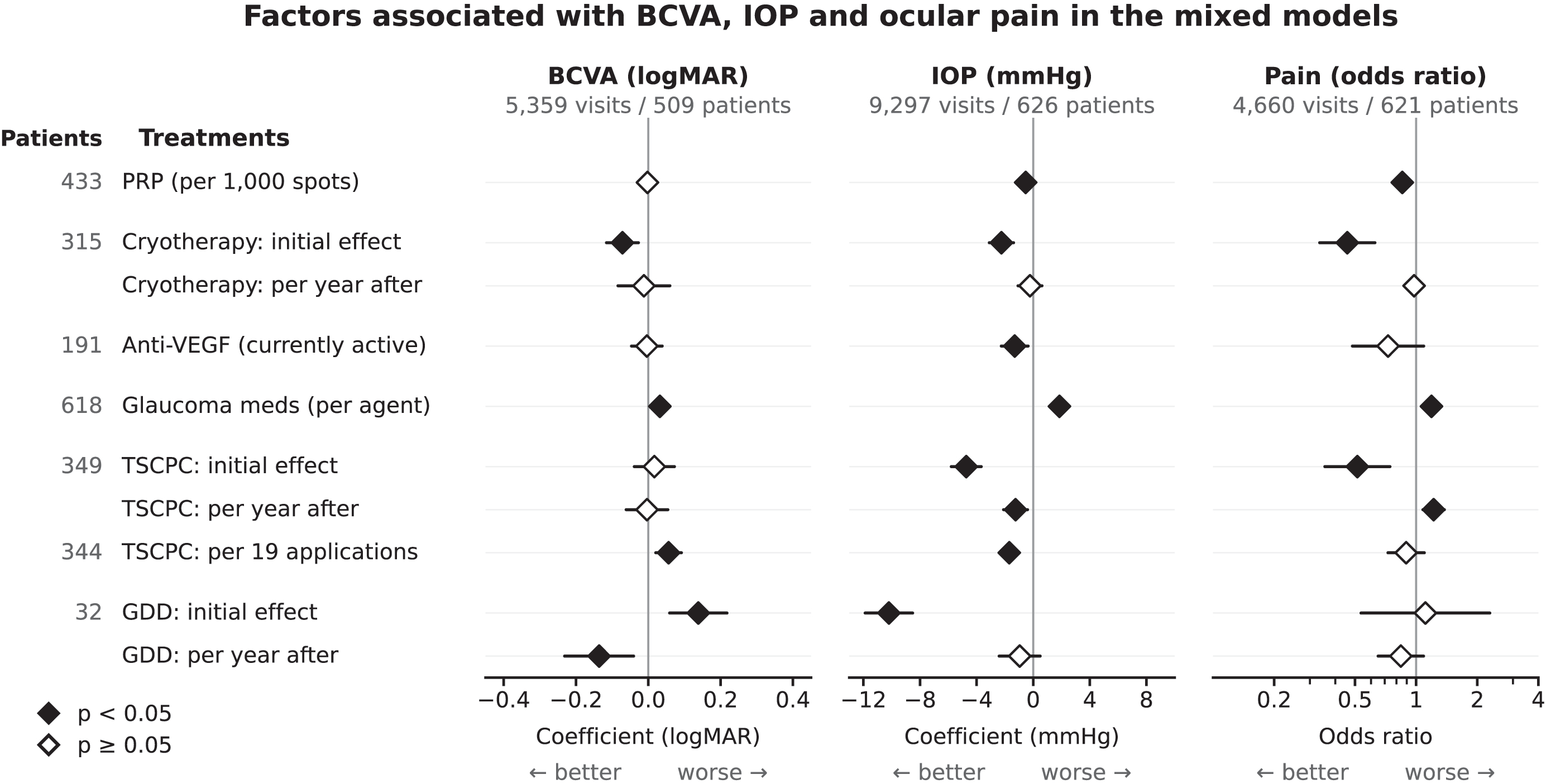
Treatment estimates after diagnosis from three mixed-effects models, each with a patient-level random effect. Procedures modelled with a step-and-slope encoding show two rows: the initial effect once the procedure has been performed, and the further change per year after it. The patient count on the left is the number contributing each treatment. Each model also included aetiology, demographic, baseline ocular, comorbidity, and other treatment covariates, shown in full in Table 2. Median TSCPC application count was 19 per session. Anti-VEGF, anti-vascular endothelial growth factor; BCVA, best-corrected visual acuity; GDD, glaucoma drainage device; IOP, intraocular pressure; PRP, pan-retinal photocoagulation; TSCPC, transscleral cyclophotocoagulation.

**Table 2.** Factors associated with BCVA, IOP and ocular pain in the mixed models.

| Factor | BCVA (logMAR) | IOP (mmHg) | Ocular pain (OR) |
| --- | --- | --- | --- |
| **Aetiology (ref. CRVO)** |  |  |  |
| Diabetic retinopathy | -0.27 (-0.40, -0.14)** | -0.9 (-3.6, 1.8) | 1.42 (0.79, 2.53) |
| CRAO | 0.01 (-0.11, 0.14) | -1.5 (-3.8, 0.9) | 1.39 (0.84, 2.29) |
| OIS | -0.09 (-0.21, 0.04) | 0.5 (-2.1, 3.1) | 0.98 (0.55, 1.77) |
| Other (grouped) | -0.07 (-0.16, 0.03) | -0.7 (-2.5, 1.0) | 1.19 (0.81, 1.76) |
| **Demographics** |  |  |  |
| Male (ref. female) | -0.04 (-0.10, 0.03) | -1.2 (-2.5, 0.1) | 0.78 (0.58, 1.04) |
| Age, per decade (ref. 75 years) | 0.00 (-0.03, 0.03) | 0.0 (-0.6, 0.6) | 0.88 (0.78, 0.99)* |
| **Lens status (ref. IOL)** |  |  |  |
| Aphakic | 0.31 (-0.01, 0.64) | 0.7 (-6.1, 7.4) | 3.62 (0.90, 14.46) |
| Cataract | 0.06 (-0.02, 0.15) | 0.2 (-1.4, 1.8) | 0.83 (0.57, 1.21) |
| Phakic, clear lens | -0.09 (-0.24, 0.06) | -2.2 (-5.4, 1.0) | 1.28 (0.62, 2.65) |
| **Baseline ocular status** |  |  |  |
| Baseline BCVA, per logMAR | 0.70 (0.65, 0.75)** | 2.4 (1.5, 3.3)** | — |
| Baseline IOP, per 10 mmHg | -0.01 (-0.04, 0.01) | 3.8 (3.3, 4.3)** | 0.97 (0.87, 1.09) |
| Angle closed | 0.13 (0.02, 0.23)* | 1.9 (-0.1, 3.9) | 1.17 (0.72, 1.92) |
| **Comorbidity and history** |  |  |  |
| Prior glaucoma | -0.04 (-0.12, 0.05) | 0.3 (-1.4, 1.9) | 0.94 (0.66, 1.35) |
| Diabetes | -0.08 (-0.16, -0.01)* | -0.3 (-1.8, 1.3) | 0.99 (0.71, 1.39) |
| Charlson comorbidity index, per point | 0.01 (-0.01, 0.03) | 0.0 (-0.4, 0.4) | 1.04 (0.95, 1.14) |
| Recent cataract surgery | -0.06 (-0.18, 0.06) | 0.7 (-2.0, 3.3) | 0.79 (0.45, 1.41) |
| Prior posterior-segment surgery | -0.01 (-0.16, 0.13) | 0.2 (-2.8, 3.1) | 1.37 (0.63, 2.97) |
| Prior PRP | 0.00 (-0.09, 0.09) | 1.8 (-0.1, 3.7) | 0.83 (0.54, 1.26) |
| Prior anti-VEGF | -0.01 (-0.11, 0.09) | -1.8 (-3.8, 0.2) | 1.39 (0.91, 2.13) |
| Prior glaucoma surgery | 0.01 (-0.11, 0.13) | -0.6 (-3.0, 1.8) | 0.95 (0.55, 1.64) |
| **Ongoing treatment** |  |  |  |
| Anti-VEGF active | 0.00 (-0.05, 0.04) | -1.3 (-2.3, -0.4)* | 0.73 (0.49, 1.09) |
| Glaucoma medications, per agent | 0.03 (0.02, 0.04)** | 1.9 (1.6, 2.1)** | 1.19 (1.10, 1.28)** |
| PRP spots, per 1000 (cumulative) | 0.00 (-0.01, 0.01) | -0.5 (-0.7, -0.4)** | 0.85 (0.81, 0.90)** |
| TSCPC applications, per 19 (cumulative) | 0.06 (0.02, 0.09)* | -1.7 (-2.3, -1.0)** | 0.89 (0.73, 1.10) |
| **Procedures and time since** |  |  |  |
| TSCPC, initial effect | 0.02 (-0.04, 0.07) | -4.7 (-5.8, -3.7)** | 0.51 (0.36, 0.74)** |
| Time since TSCPC, per year | 0.00 (-0.06, 0.05) | -1.2 (-2.1, -0.4)* | 1.22 (1.08, 1.38)* |
| GDD, initial effect | 0.14 (0.06, 0.22)** | -10.2 (-11.9, -8.5)** | 1.11 (0.54, 2.30) |
| Time since GDD, per year | -0.14 (-0.23, -0.04)* | -1.0 (-2.4, 0.5) | 0.84 (0.65, 1.09) |
| Cryotherapy, initial effect | -0.07 (-0.12, -0.03)* | -2.2 (-3.1, -1.4)** | 0.46 (0.34, 0.63)** |
| Time since cryotherapy, per year | -0.01 (-0.08, 0.06) | -0.2 (-1.1, 0.6) | 0.98 (0.87, 1.09) |
| Vitrectomy, initial effect, any time | -0.05 (-0.16, 0.05) | -2.4 (-4.6, -0.3)* | 0.79 (0.38, 1.63) |
| **Endarterectomy** |  |  |  |
| In OIS eyes | 0.15 (-0.01, 0.31) | 7.7 (4.1, 11.2)** | 3.83 (1.38, 10.66)* |
| Difference, OIS vs non-OIS | 0.32 (0.03, 0.60)* | 3.5 (-2.2, 9.1) | 4.10 (0.93, 18.16) |

Estimates come from three mixed-effects models, each with a patient-level random effect: BCVA (5,359 visits, 509 patients), IOP (9,297 visits, 626 patients), and pain (4,660 visits, 621 patients). BCVA and IOP columns show regression coefficients (positive BCVA = worse vision, positive IOP = higher pressure); the pain column shows odds ratios. Baseline BCVA was not entered into the pain model ("—"). Cumulative PRP spots are expressed per 1000, and cumulative TSCPC applications per 19 (the median among treated eyes). Estimates are associational, not causal. Values are estimate (95% CI). \*p<0.05, \*\*p<0.01. Anti-VEGF, anti-vascular endothelial growth factor; BCVA, best-corrected visual acuity; CI, confidence interval; CRAO, central retinal artery occlusion; CRVO, central retinal vein occlusion; DR, diabetic retinopathy; GDD, glaucoma drainage device; IOL, intraocular lens; IOP, intraocular pressure; OIS, ocular ischaemic syndrome; OR, odds ratio; PRP, pan-retinal photocoagulation; TSCPC, transscleral cyclophotocoagulation.

Interaction tests examined whether treatment associations with BCVA and IOP varied by aetiology, angle, or baseline severity. Baseline-severity interactions appeared throughout but likely reflected the greater room for improvement in the most impaired eyes, so were not pursued. With separate time courses per group, TSCPC (−5.2 mmHg, 95% CI −7.2 to −3.3) and cryotherapy (−3.2 mmHg, 95% CI −5.3 to −1.0) were associated with greater IOP lowering in closed-angle than open-angle eyes (Supplementary Table 3). In DR compared with CRVO, cryotherapy was associated with better BCVA (−0.25 logMAR, 95% CI −0.35 to −0.15) but higher IOP (3.2 mmHg, 95% CI 1.2–5.3), and PRP with higher IOP (per 1000 spots, 0.5 mmHg, 95% CI 0.1–0.9; Supplementary Table 3). The angle finding held with angle-specific baseline IOP.

Primary findings were robust across most sensitivity analyses (Supplementary Table 4). However, in the ordinal BCVA model, the GDD and cryotherapy associations lost significance but kept their direction. All covariate variance inflation factors remained below 4, excluding the time spline basis.

### Machine learning

The cross-validation mean RMSE was 0.34 logMAR (standard deviation, 0.01) for BCVA and 9.6 mmHg (standard deviation, 0.4) for IOP. Cross-validation RMSE exceeded training RMSE by 0.05 logMAR and 2.5 mmHg. In the model including the previous visit’s BCVA and IOP, these features had the highest SHAP values. Without them, baseline BCVA and IOP ranked highest. The ML results were broadly consistent with the LMM. In the BCVA model, a more closed angle was associated with worse vision and DR with better vision. In the IOP model, corneal oedema, pain, and more glaucoma medications were associated with higher IOP. Time since diagnosis ranked highly in both analyses (Figure 2). Analysing treatments separately to reveal rarer features not surfaced in the main SHAP summary also showed similar results to the LMM (Supplementary Table 5).

**Figure 2.**
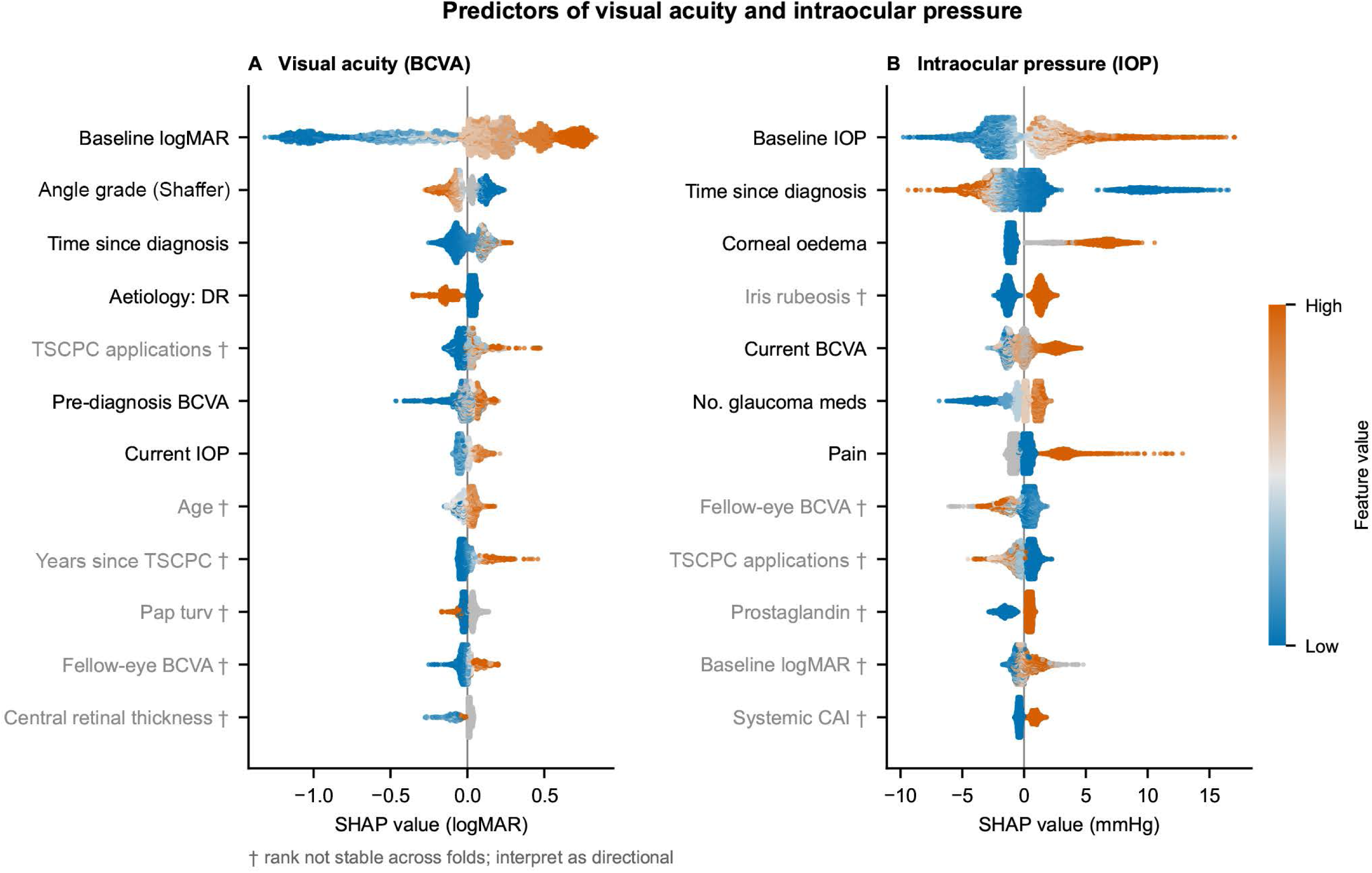
SHAP summary of the gradient-boosted models for BCVA (Panel A) and IOP (Panel B). Each dot is one visit, positioned by its SHAP value: the amount that variable moved the model’s prediction above or below the average, in the units of the outcome. Colour shows the value of the variable itself, from low (blue) to high (orange). A row where orange dots sit mainly on one side of zero indicates the direction of the association. Variables are ordered by mean absolute SHAP value, and those marked † did not hold a stable rank across cross-validation folds. These models were exploratory, and SHAP values describe how the model uses each variable rather than any causal effect. BCVA, best-corrected visual acuity; DR, diabetic retinopathy; IOP, intraocular pressure; SHAP, Shapley Additive exPlanations; TSCPC, transscleral cyclophotocoagulation.

To examine the shape of the highest-ranked features, we used SHAP dependence plots (Figure 3). These plots revealed that an open angle of Shaffer grade above 1 was associated with worse predicted BCVA in eyes with good baseline BCVA compared to poor baseline BCVA (including NLP). Plotting by cumulative TSCPC applications showed that the transition from none to the first applications contributed the largest reduction in predicted IOP, with little additional reduction at further sessions.

**Figure 3.**
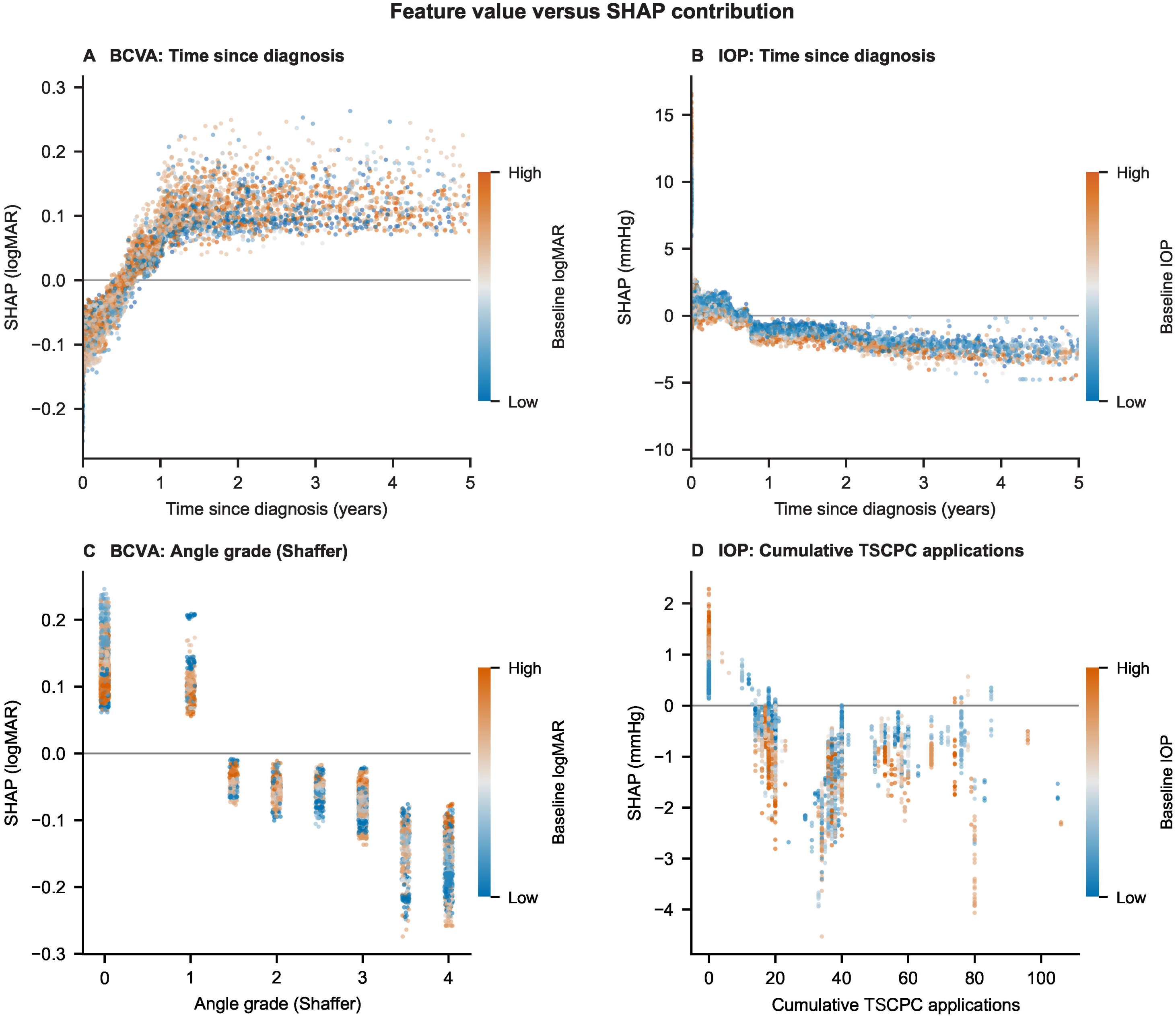
SHAP dependence plots for selected variables in the gradient-boosted BCVA and IOP models. Each panel plots one variable’s value against its SHAP contribution across all visits. Panel A, time since diagnosis for BCVA: the contribution rises over the first year, then plateaus. Panel B, time since diagnosis for IOP: the contribution falls over the first year and continues to decline, with higher baseline IOP showing the greater reduction. Panel C, Shaffer grade for BCVA: more open angles carry an increasingly negative contribution. Panel D, cumulative TSCPC applications for IOP: the contribution is positive at zero applications, drops once applications accumulate, and flattens at higher counts. These SHAP values describe how the exploratory models use each variable rather than any causal effect. BCVA, best-corrected visual acuity; IOP, intraocular pressure; SHAP, Shapley Additive exPlanations; TSCPC, transscleral cyclophotocoagulation.

The ML analysis identified a Shaffer grade-DR interaction. We tested it in the LMM, where it did not reach statistical significance (−0.04 logMAR per Shaffer grade, 95% CI −0.09 to 0.01, *p*=0.151). In a broad SHAP interaction screen, after accounting for baseline severity, no treatment was more strongly associated with better BCVA or lower IOP in any particular type of eye.

The results were stable across the learning-rate and depth sensitivity analyses. Without the previous measurements, neither baseline nor biomicroscopy features contributed much to predictive performance (Supplementary ML analyses).

## Discussion

We aimed to analyse the factors associated with BCVA, IOP, and pain outcomes in NVG using two approaches: a primary LMM analysis with pre-specified covariates and an exploratory ML analysis that searched the full dataset. The models showed similar results, with no novel clinical findings arising from the fuller ML dataset.

Baseline characteristics carried most of the visual prognosis, whereas treatments were associated mainly with lower IOP and reduced pain rather than better vision. Among the baseline factors, age, sex, and CCI were not associated with BCVA or IOP. None appeared in the main SHAP summary. Some studies reported young age as a risk factor for operative failure^9^ ^36^ ^37^, but others did not^5^ ^8^. Our findings agree with the latter. Sex has not been linked to glaucoma outcomes^38^. Angle closure at baseline has been linked to poorer visual outcomes^39^ as our study found. Of the marked aetiology differences reported in this cohort previously^13^, only better BCVA in DR persisted as an independent association after adjustment for baseline severity, ocular status, age, and time since diagnosis. DR was also the only aetiology feature to surface in the BCVA SHAP summary.

Cryotherapy and TSCPC have been found to lower IOP whereas visual outcomes still deteriorated^40^ ^41^. We showed that cryotherapy was associated with better BCVA in the continuous LMM, although this did not hold in the ordinal model. The variation came from the coarser outcome, not the random-effects structure: both stayed significant when the continuous model used random intercepts alone. Additionally, both cryotherapy and TSCPC were associated with lower IOP, particularly in closed-angle eyes. The effect is consistent with cyclophotocoagulation reducing aqueous production independently of the angle^3^, though the same pattern for cryotherapy is unexplained. The modest, short-lived IOP association of anti-VEGF, and its absence of any BCVA or pain signal, fits evidence that a single injection lowers pressure only briefly and does not treat established NVG alone^42^ ^43^.

Pain has been researched less thoroughly in NVG, but the literature supports cryotherapy and TSCPC reducing pain^6^ ^44^ ^45^. Our models showed that the initial effects of cryotherapy, TSCPC, and each 1000 spots of PRP were associated with lower pain odds. GDDs were associated with a sustained IOP reduction and, after an immediate worsening, with better BCVA over subsequent years, supported by the literature^4^ ^46^. The small, highly selected GDD subgroup and the likelihood of recovery from peri-operative worse BCVA warrant caution in interpreting the results.

The LMMs and the ML rest on different assumptions, so shared associations are more credible. Mostly stable feature rankings and a small train-validation gap suggest the SHAP attributions are not overfitting artefacts. Previous studies reported that a second TSCPC treatment retains IOP-lowering efficacy comparable to the first^47^ ^48^. Our ML showed otherwise: the first session contributed most to predicted IOP. Eyes needing repeat treatment are likely refractory to it.

More glaucoma medications were associated with worse vision, higher IOP, and more pain. The association illustrates confounding by indication, a central limitation of retrospective treatment analyses that applies throughout this study. Other limitations include bias at data collection, which was previously addressed^13^; sicker eyes being seen more often, accentuating severe trajectories; competing endpoints employing informative censoring, such as morbidity and mortality; the NLP absorbing state in the primary BCVA LMM; and the single-centre nature limiting the results.

## Conclusion

Vision in this cohort was largely set by the state of the eye at diagnosis. Treatments mostly related to lower IOP and less pain rather than to better vision. Pressure control and pain relief therefore remain realistic goals even when sight cannot be saved. Peripheral retinal cryotherapy stood out, linked to both lower IOP and less pain, a pattern seldom reported in neovascular glaucoma. Clinicians allocated treatment by judgement, so confounding by indication may bias these associations in either direction.

## Data Availability

All data produced in the present work are contained in the manuscript and supplementary data.

## Declaration Statements

## Acknowledgements

In preparing this work, the lead author (GS) used Claude Opus 4 and 5 (Anthropic, San Francisco, CA, USA)^25^ during 2026. This tool was used to improve the cohesion and coherence of the language. AI use in drafting and debugging the analysis code is described in the Methods. The tool was not used to design the study, select statistical methods, analyse data, or interpret findings. GS had full access to all the data in the study and takes responsibility for the integrity of the data and the accuracy of the data analysis. The lead author would like to thank the Biostatistics Consulting Service provided by the Biostatistics Unit, Faculty of Medicine, University of Helsinki, for helping plan the mixed-effects models and Rami Luisto, Faculty of Information Technology, University of Jyväskylä, who assisted in planning the machine learning approach.

## Contributors

GS: Conceptualization, Data curation, Funding Acquisition, Formal analysis, Investigation, Methodology, Software, Validation, Visualization, Writing – original draft. MvF: Data curation. AD: Conceptualization, Methodology, Writing – review & editing. VV: Conceptualization, Methodology, Supervision, Writing – review & editing. PS: Conceptualization, Supervision, Writing – review & editing. MH: Conceptualization, Funding Acquisition, Methodology, Supervision, Writing – review & editing.

