## Supplementary for "Predictors of Visual Acuity, Intraocular Pressure, and Pain in Neovascular Glaucoma: A Mixed-Effects Model and Machine Learning Cohort Analysis"

### Supplements

#### Supplementary Methods

##### Inclusion criteria

The IOP threshold restricted the cohort to eyes requiring treatment and was similar to thresholds used previously^1 2^.

##### Visual acuity

Chart acuities were converted to logMAR as −log₁₀(decimal acuity), and Early Treatment Diabetic Retinopathy Study letter scores as 1.7−0.02·letters^3^. Partly read lines were rounded down to the last full line and plus notations were ignored. Counting fingers and hand motion were assigned bounded logMAR intervals based on testing distance rather than single values. Values of 2.00 logMAR for counting fingers and 2.30 logMAR for hand movement at a standard testing distance of 0.30 m were used^4^. A half-width of 0.10 logMAR, one chart line, was applied to each acuity. Recorded testing distance was mapped within each interval by interpolation on log₁₀ distance, pinned so that 0.30 m returns the reference value, with 0.10 m and 3.00 m returning the interval bounds. Distances outside this range were clamped, and 'ad oculum' was treated as 0.10 m. Where no distance was recorded, the 0.30 m reference value was applied. Light perception and no light perception were assigned 2.70 and 3.00 logMAR^5^.

##### Intraocular pressure

IOP was recorded before pharmacological mydriasis whenever possible. A small number of measurements taken by methods other than Goldmann applanation or rebound tonometry were pooled with the rebound values.

##### Pain

Pain status was categorised as absent, mild, or severe. Only ocular pain or aching was included; other non-specific sensations were excluded.

##### Biomicroscopic grading

**Anterior chamber angle.** The most open angle recorded at a visit was graded on the Shaffer scale^6^, from grade 0 (closed) to grade 4 (fully open), and could take half-grades. Where the notes stated only that the angle was open, we assigned grade 3. Angle width was read without indentation.

**Peripheral anterior synechiae.** Synechial extent was categorised as mild for 0–90°, moderate for 90–270° and severe for 270–360°. Extents falling exactly at the 90° and 270° boundaries were counted as moderate. When extent was not documented we assigned the moderate category.

**Anterior chamber.** Cells were graded by the Standardization of Uveitis Nomenclature scheme ^7^. Chamber depth was classified as shallow or as normal-to-deep. Hyphema was recorded as present or absent.

**Cornea and conjunctiva.** We recorded conjunctival hyperaemia and, for the cornea, oedema, Descemet membrane folds and neovascularisation.

**Iris.** Rubeosis and vascular congestion were recorded separately. An eye scored as congested was also scored as not having rubeosis.

**Lens.** Lens status was recorded as phakic without cataract, cataract, pseudophakia or aphakia. Pseudoexfoliation was noted separately.

**Vitreous, optic disc and retina.** We recorded vitreous haemorrhage, and at the optic disc, oedema and neovascularisation. The macula and the peripheral retina were assessed separately for haemorrhage, neovascularisation, oedema, and ischaemia.

##### Optical coherence tomography and visual fields

Central retinal thickness was taken from SPECTRALIS macular scans (Heidelberg Engineering GmbH, Heidelberg, Germany) using the Heidelberg Eye Explorer software. Scans with a quality score below 15, or without successful automated segmentation, were discarded. Visual field data, from both Humphrey automated and Goldmann kinetic perimetry, were too sparse after diagnosis to support longitudinal analysis.

##### Charlson comorbidity index

The Charlson comorbidity index (CCI) was calculated for each patient from the imported full hospital diagnosis lists. Primary care diagnoses were not included. The register was built using the ICD-10 codes of Ludvigsson *et al.*^8^ with weights of Charlson *et al.*^9^.

##### Linear mixed-effects models

A sensitivity analysis for BCVA used a cumulative link mixed-effects model using ordinal categories (WHO ICD-11 classification^10^: decimal acuity ≥0.33, mild impairment or better, 0.33 to 0.1, moderate visual impairment and worse than 0.1, severe visual impairment or worse). IOP sensitivity analyses excluded hypotony cases (IOP <5 mmHg, 3% of visits in 13% of patients) and used only Goldmann applanation tonometry. Pain was checked by treating all visits without pain data as pain-free. Sensitivity analyses using only baseline values from the diagnosis date, instead of within 30 days, was also computed.

##### Machine learning

Refraction was excluded from the models. It is recorded only for eyes seeing counting fingers or better, so its absence carries outcome information, and its apparent predictive value reflects that recording pattern rather than any refractive signal.

The number of estimators was fixed at 500 in advance. We inspected the learning curves afterwards to confirm this was sufficient, with training and validation loss having plateaued, and did not tune the value.

#### Supplementary Tables

##### Supplementary Table 1.

| Modelling hyperparameters | Value | Purpose |
| --- | --- | --- |
| n_estimators | 500 | Boosting rounds, fixed with no early stopping |
| max_depth | 4 | Shallow trees, limiting interaction order |
| learning_rate | 0.05 | Slow learning to average out noise |
| min_child_weight | 10 | Minimum rows per leaf, blocking single-patient leaves |
| gamma | 1 | Minimum loss reduction required to split |
| subsample | 0.8 | Row subsampling per tree |
| colsample_bytree | 0.8 | Feature subsampling per tree |
| reg_lambda | 1.0 | L2 penalty on leaf weights |
| reg_alpha | 0 (default) | L1 penalty, left at default |
| Fixed operational settings |  |  |
| objective | reg:squarederror | Squared-error loss for the continuous outcomes |
| tree_method | hist | Histogram-based split finding |
| random_state | 42 | Fixed seed for subsample and colsample draws |

The same specification was applied to the BCVA and IOP models. Hyperparameters were fixed in advance rather than tuned. No early stopping was used. Validation was 5-fold patient-level GroupKFold with no held-out test set. Hyperparameters not listed used XGBoost 2.0.3 defaults. The sensitivity analysis varied max_depth (3 and 5) and learning_rate (0.03 and 0.1) one parameter at a time, with all other values held at the table above.

##### Supplementary Table 2.

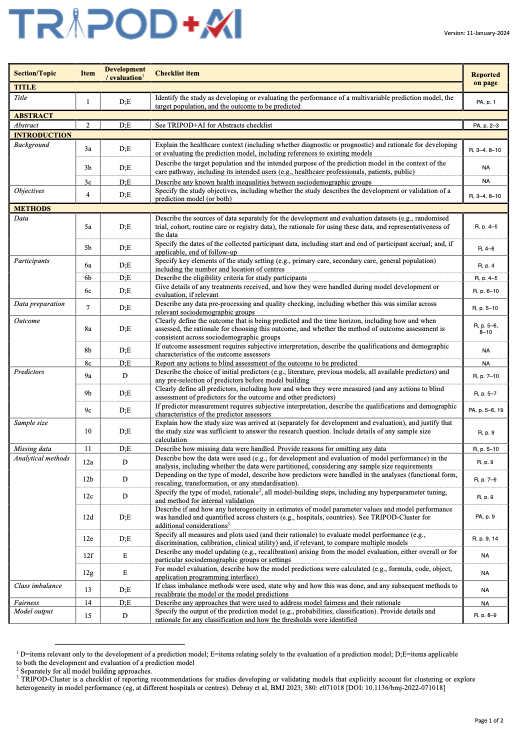

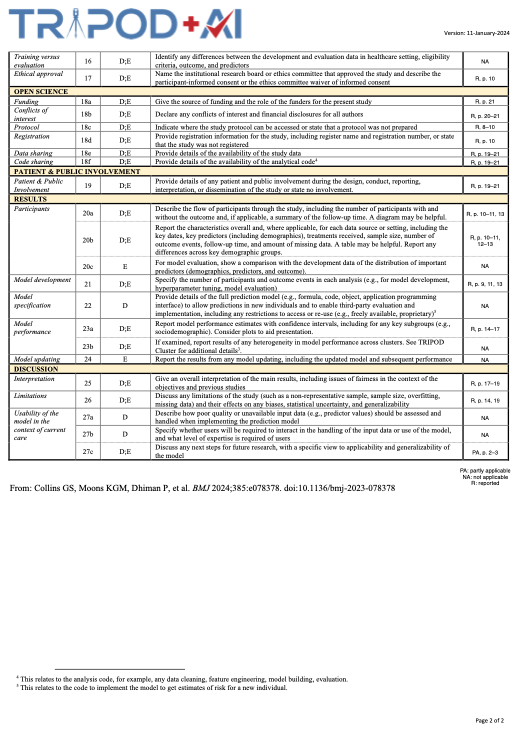

##### Supplementary Table 3.

| **Supplementary Table 3. Treatment associations that varied by angle or aetiology.** | | | | |
| --- | --- | --- | --- | --- |
| **Outcome** | **Treatment** | **Comparison** | **Estimate (95% CI)** | ***p*** |
| **By anterior chamber angle** | |  |  |  |
| IOP, mmHg | TSCPC | Closed vs open angle | −5.2 (−7.2, −3.3) | <0.001 |
| IOP, mmHg | Cryotherapy | Closed vs open angle | −3.2 (−5.3, −1.0) | 0.004 |
| **By aetiology** |  |  |  |  |
| BCVA, logMAR | Cryotherapy | DR vs CRVO | −0.27 (−0.37, −0.16) | <0.001 |
| IOP, mmHg | Cryotherapy | DR vs CRVO | 3.3 (1.2, 5.3) | 0.002 |
| IOP, mmHg | PRP | DR vs CRVO | 0.5 (0.1, 0.9) | 0.022 |

##### Supplementary Table 4.

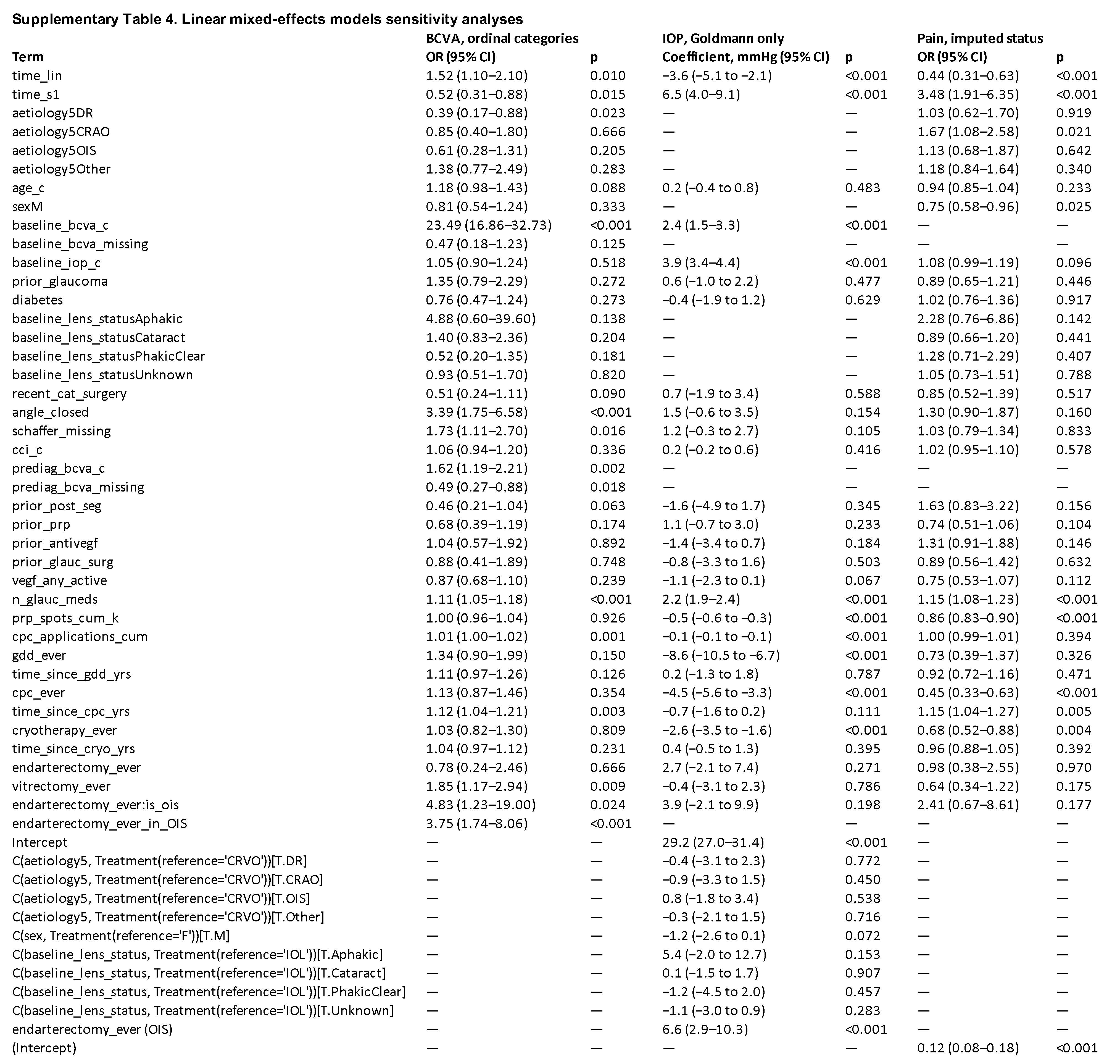

##### Supplementary Table 5.

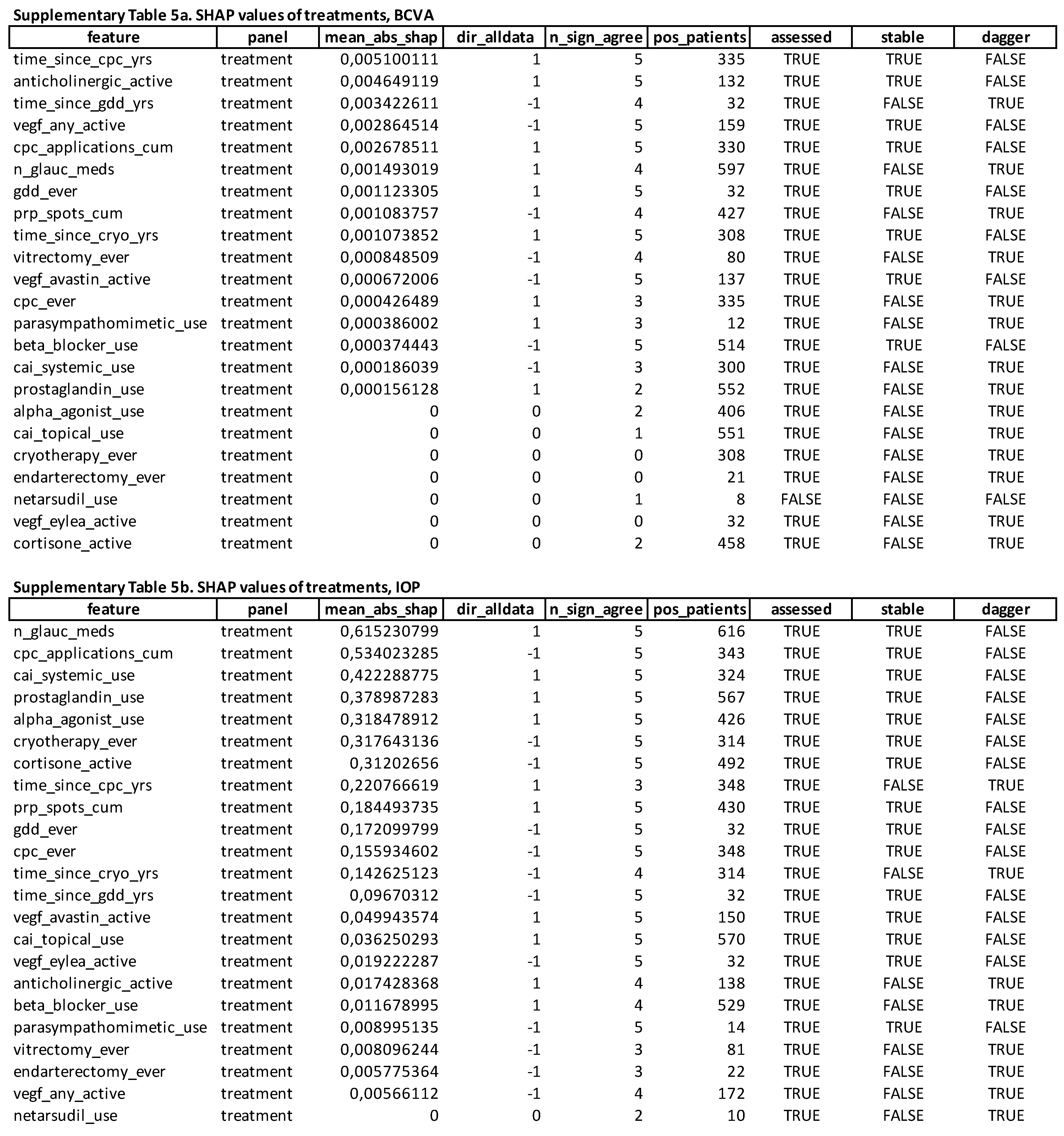

##### Supplementary ML analyses

The sensitivity analysis varying learning_rate and depth showed similar results to the primary model, with a spread of 0.01 logMAR and 0.6 mmHg across iterations. Examining cross-validation RMSE for the BCVA model without the previous measurements showed an increase in RMSE of 0.21 logMAR, from 0.36 (standard deviation, SD, 0.01) to 0.57 (SD, 0.03). Comparing the model without the previous measurements to a model without any biomicroscopic information only increased RMSE by 0.03, to 0.60 (SD, 0.03). Only including baseline values resulted in an RMSE of 0.65 (SD, 0.02). The IOP model showed the same ordering: the lag contributed 1.8 mmHg, biomicroscopy 0.2 mmHg with the lag present and 0.5 mmHg without it.

10. World Health Organization. World report on vision, 2019.
